# Childhood inductive reasoning, cardiovascular and cardiometabolic morbidity and dementia risk – a population representative cohort study across six decades

**DOI:** 10.64898/2026.05.11.26352876

**Authors:** Kristine B. Walhovd, Anne Ingeborg Berg, Sandra Buratti, Jonas Burén, Pär Bjälkebring, Martin Fischer, Isabelle Hansson, Linda Hassing, Anna-Carin Jonsson, Lina Jonsson, Magnus Lindwall, Therese Nilsson, Andreas Segerberg, Valgeir Thorvaldsson, Mikael Landén, Alli Klapp, Martin Lövdén

## Abstract

**Background:** Higher cognitive ability in late adolescence and young adulthood associate with reduced risk of dementia, but such assessments are influenced by educational selection. Whether specific cognitive abilities earlier in childhood associate with later dementia risk, independent of known associations with cardiovascular disease (CVD) and diabetes, remains unclear.

**Methods:** We studied a Swedish population-representative birth cohort with cognitive testing at age□13 (n□=□10,539 born in 1948). Dementia and somatic morbidity were ascertained from nationwide inpatient and cause-of-death registers through November 2025 (>□6 decades). Cox models estimated associations between childhood inductive reasoning, verbal and spatial ability scores and dementia, somatic morbidity, CVD and diabetes, each modelled as outcomes and as time-varying covariates in dementia models.

**Results:** During follow-up, 287 individuals (2.7%) developed dementia. Higher childhood inductive reasoning associated with lower risk of dementia (HR per SD□=□0.84, 95%□CI 0.72–0.98), somatic morbidity, CVD (HR□0.88, 95%□CI 0.83–0.92), and diabetes (HR□0.74, 95%□CI 0.64–0.86). Verbal and spatial abilities were not independently associated with dementia. Somatic morbidity, CVD, and diabetes associated with dementia risk (HRs ∼1.63–2.74), but only modestly attenuated the inductive reasoning–dementia association (HRs ∼0.84–0.85). Findings replicated in an expanded cohort including individuals born in 1953 (total n□=□19,919) and were robust to adjustment for parental and midlife education.

**Conclusions:** Higher childhood inductive reasoning was associated with lower dementia risk across six decades, and this relationship was not substantially attenuated by adjustment for education or clinically manifest cardiovascular or cardiometabolic disease. These findings are consistent with a life-course perspective in which early neurodevelopmental characteristics contribute to dementia risk alongside major vascular and metabolic risk factors.

**Funding:** Riksbankens Jubileumsfond (M23-0040); Marks Guest Professorate fellowship.

## Background

Individual differences in cognitive function emerge early in development and are associated with a wide range of health and functional outcomes across the lifespan (1). In later life, higher cognitive ability assessed in adolescence and young adulthood has been linked to reduced risk of dementia in large population-based registry studies (2–5). However, cognitive performance in late adolescence and early adulthood is increasingly shaped by schooling and family environment (6, 7), including parental education. As a result, it remains difficult to disentangle whether associations between early cognitive performance and later dementia risk reflect early emerging cognitive vulnerability or downstream influences related to education and adolescent experiences.

Evidence from the Scottish Mental Survey of 1932/1947 suggests the relevant time window may be in childhood, linking higher cognitive ability at age 11 to lower dementia risk (2, 3). Notably, especially strong protective associations among women were reported(4). The largest registry-based investigations, however, rely on cognitive testing at mandatory military conscription,(5, 8, 9) which is limited to men and conducted at an age when educational and social influence is already considerable. Together, these findings raise the question of whether variation in childhood cognitive ability, before major divergence in educational pathways and other environmental influences, is associated with dementia risk later in life.

Importantly, most evidence linking early cognition to dementia risk is based on global intelligence scores. Little is known about whether specific cognitive domains in childhood have differential predictive value. If so, this would move the field beyond a generic global cognition view of protection and towards a more nuanced, domain-specific model. Domain-specific cognitive patterns can point to distinct neural and developmental factors linking early cognition to later dementia and could in principle guide the design of early preventative programs. Huang et al.(10) provided some evidence for this, showing associations of lower mechanical reasoning in men and lower verbal memory in women with risk of dementia linking U.S. high-school aptitude subtests to Medicare records, but did not assess possible associations with e.g. other diseases in adulthood. Understanding which early-life cognitive abilities matter most, may help shift prevention efforts toward a life-course perspective, recognizing childhood as a meaningful window for long-term brain health.

Moreover, as cognitive ability at young age has been linked to multiple diseases associated with dementia risk, especially cardiovascular (CVD) and cardiometabolic diseases (diabetes) (1, 11–14), it is important to assess to what extent adjustment for such diseases may attenuate any relationship between domain-specific childhood cognitive functioning and dementia. Lower intelligence in childhood has been firmly associated with increased risk of cardiovascular events in adulthood, even after adjustment for confounding variables(12, 14–17). Similarly, lower cognitive function at young age has been linked to risk of premature diabetes in adulthood (14). However, it is unknown whether possible associations of specific childhood cognitive domains and dementia risk may be based on increased susceptibility to such other diseases, or whether independent relationships can be identified.

To address these knowledge gaps, we analyze the Swedish 1948 UGU birth cohort, in which population representative participants completed tests of inductive reasoning, verbal ability, and spatial problem-solving at age 13 (18, 19). By linking primary school stage cognition to dementia diagnoses as well as major somatic morbidity, CVD and diabetes in nationally representative population registries (20, 21) with follow-up spanning more than six decades, our objectives are to test whether specific cognitive domains are uniquely predictive of dementia independently of pre-test educational differences, and when adjusting for other major diseases. We deliberately focus on inpatient morbidity and mortality-registered dementia to maximize population representativity and minimize bias related to healthcare access, educational level, and diagnostic intensity (20). Parental education and prospective confounding by differences in individualś own educational attainment in midlife were corrected for in analyses.

## Methods

### Study population

The UGU study is a longitudinal Swedish school-based cohort (18, 19). The 1948 birth cohort analyzed here, included Swedish children born on 5th, 15^th^ or 25^th^ of the month nationwide (see Figure S1 for a flowchart showing the full sampling frame) and completed cognitive testing in 6th grade in 1961 (n = 10,539; 5372 men, 5167 women). During follow-up (May 1961-Nov 2025), 287 participants were diagnosed with dementia in the National Patient Register Inpatient Register (NPR-IPR) and/or Cause of Death Register (CDR). The sample characteristics by dementia status is shown in Table 1. Parental education was recorded at testing; participants’ educational attainment was obtained from the 1990 Census (age ≈ 42; n = 9640) (Supplement, Table S1).

**Table 1.** Characteristics of the 1948 Birth Cohort According to Dementia Status During Follow-up. Values are presented as n (%) for categorical variables, mean (SD) for continuous cognitive and education measures, and median (interquartile range [IQR]) for age variables. Parental education was recorded as a school administrative variable at the time of testing and coded on an ascending scale from 1 to 4. Midlife education was obtained from the 1990 Swedish Census, coded on a scale from 1 to 7, and was missing for 899 participants (see Table S1).

| Characteristic | No Dementia<br>(n=10,252) | Dementia<br>(n=287) |
| --- | --- | --- |
| Female, n (%) | 5037 (49.1%) | 130 (45.3%) |
| Inductive reasoning, mean (SD) | 19.36 (7.96) | 18.32 (7.96) |
| Spatial ability, mean (SD) | 21.06 (7.01) | 20.70 (7.50) |
| Verbal ability, mean (SD) | 22.32 (7.01) | 21.90 (7.11) |
| Parental education, mean (SD) | 1.23 (0.54) | 1.24 (0.55) |
| Midlife education, mean (SD) | 3.38 (1.71) | 3.34 (1.77) |
| Age at dementia diagnosis, median (IQR), y | — | 73.57 (5.98) |

### Cognitive tests

Verbal, spatial and inductive reasoning abilities were tested with 1) antonyms (choosing the opposite of a word from four options), 2) metal folding (determine which of four figures you get if folding a pictured “sheet metal piece”), and 3) number series (continuing a number-series, where six numbers are given, with two more numbers) (22). Test scores had acceptable reliabilities (r = .87-.92) and correlated within expected range (r = .42-.56, see Supplement). Score distributions were approximately normal (Figure S2).

### Dementia and other disease outcome coding

Data were retrieved from registries until November 3^rd^ 2025. Dementia was identified from the NPR-IPR and CDR (starting 1964 and 1952, respectively) using ICD-9-10 codes. A participant was classified as having dementia if any hospitalization or death record contained a relevant code; date of diagnosis was defined as the earliest recorded event. Major somatic morbidity, cardiovascular disease (CVD), and diabetes were identified from NPR-IPR using ICD-7–10 codes (Supplement). To examine whether associations between childhood cognition and dementia risk were attenuated by adjustment for broader susceptibility to medical morbidity, we included diagnoses from an index of overall somatic major morbidity based on the Charlson Comorbidity Index (CCI) (23–25). We identified the earliest recorded diagnosis, prior to or concurrent with any diagnosis of dementia, corresponding to any Charlson condition other than dementia. Likewise, we identified CVD, including cerebrovascular disease, myocardial infarction, and heart failure, as well as diabetes, defined as the earliest inpatient diagnosis prior to or concurrent with any diagnosis of dementia (see Supplementary Information).

In the 1948 cohort, data were received for a total for 11,945 persons that could be coupled to registries. Nine of these persons opted out of further registry linkage, leaving 11,936 individuals, including a total of 329 dementia cases, i.e. 2.75%. 10,539 of these persons had all initial predictor variables recorded (three cognitive measures, parental education and sex), of whom 287 (2.72%) had a dementia diagnosis at end of follow-up (see Figure S1, Table 1). In the analytic sample (n = 10,539), 5,183 participants were hospitalized at least once with a major somatic morbidity condition during follow-up, of whom 2,367 had CVD, and 308 had diabetes (for sample characteristics by disease groups, see Supplement, Tables S2-S4).

Registration in the CDR and NPR-IPR has limited sensitivity, with diagnoses typically recorded later than in dedicated longitudinal clinical assessments but has high specificity and positive predictive value (21). While seeking specialized healthcare in out-patient services is selective to socioeconomic status, even adjusted for health needs, hospitalizations (registered in NPR-IPR) are less selective (20) and CDR contains all deaths with main and concomitant causes. By retrieving dementia diagnoses from CDR and NPR-IPR, and other diagnoses from NPR-IPR, we aim for the least selective, most population representative, outcome measures.

### Statistical analyses

Analyses were performed in R v4.3.3. Cox proportional hazards models with age as the underlying timescale were used to estimate hazard ratios (HRs). All continuous predictors were standardized. Cognitive predictors were entered jointly and compared with a reduced model using likelihood ratio tests. Major somatic morbidity, cardiovascular disease (CVD), and diabetes were modelled both as outcomes (first incident event) and as time-varying covariates in dementia models. Time-varying covariates were constructed (using the tmerge function, survival package), so that each condition switched from 0 to 1 at the age of first diagnosis, ensuring only morbidity preceding dementia onset was counted. Because death before dementia onset is a competing risk, Fine–Gray sub-distribution hazard models were estimated as a sensitivity analysis (cmprsk package). Additional sensitivity analyses were also carried out to address differences in subtest reliability, birth month and possible loss to follow-up by emigration as potential sources of bias (Supplementary statistical analyses and Supplementary Results). Cox models with age as the timescale implicitly adjust for age at event and properly handle the right-censored, time-to-event structure. Non-linearity was evaluated using generalized additive models (mgcv package, Supplementary statistical analyses) using a logistic link on a binary dementia outcome: these models are used as a robustness check and are not directly comparable to the Cox models. To assess multicollinearity among predictors, variance inflation factors (VIFs) were computed for all variables in the primary dementia model (car package). All VIFs were below 2·0 (range 1·02–1·62), indicating no problematic collinearity.

The funders had no role in study design, data collection, data analysis, data interpretation, or writing of the report. The corresponding author had full access to all data in the study and had final responsibility for the decision to submit for publication.

## Results

We first compared a reduced Cox model including only covariates with a full model including all three cognitive measures. The likelihood ratio test indicated that the cognitive predictors as a group contributed significantly to dementia prediction as a set (Δχ²(3) = 9.056, p = 0.029).

In Cox proportional hazards models including all cognitive measures, sex, and parental education, with age as the underlying time scale, inductive reasoning was the only cognitive score uniquely and significantly associated with dementia risk (Figure 1A, Table S5). One SD higher inductive reasoning score was associated with a 16% lower risk (HR = 0.84, 95% CI: 0.72–0.98). Neither the spatial (HR = 0.99, 95% CI: 0.86–1.13) nor the verbal (HR = 1.00, 95% CI: 0.86–1.15) subtest showed significant associations. Parental education was not associated with dementia risk (HR = 1.04, 95% CI: 0.93–1.17). Sex showed a significant association, with women having a lower hazard than men (HR = 0.79, 95% CI: 0.62–0.99).

**Figure 1:**
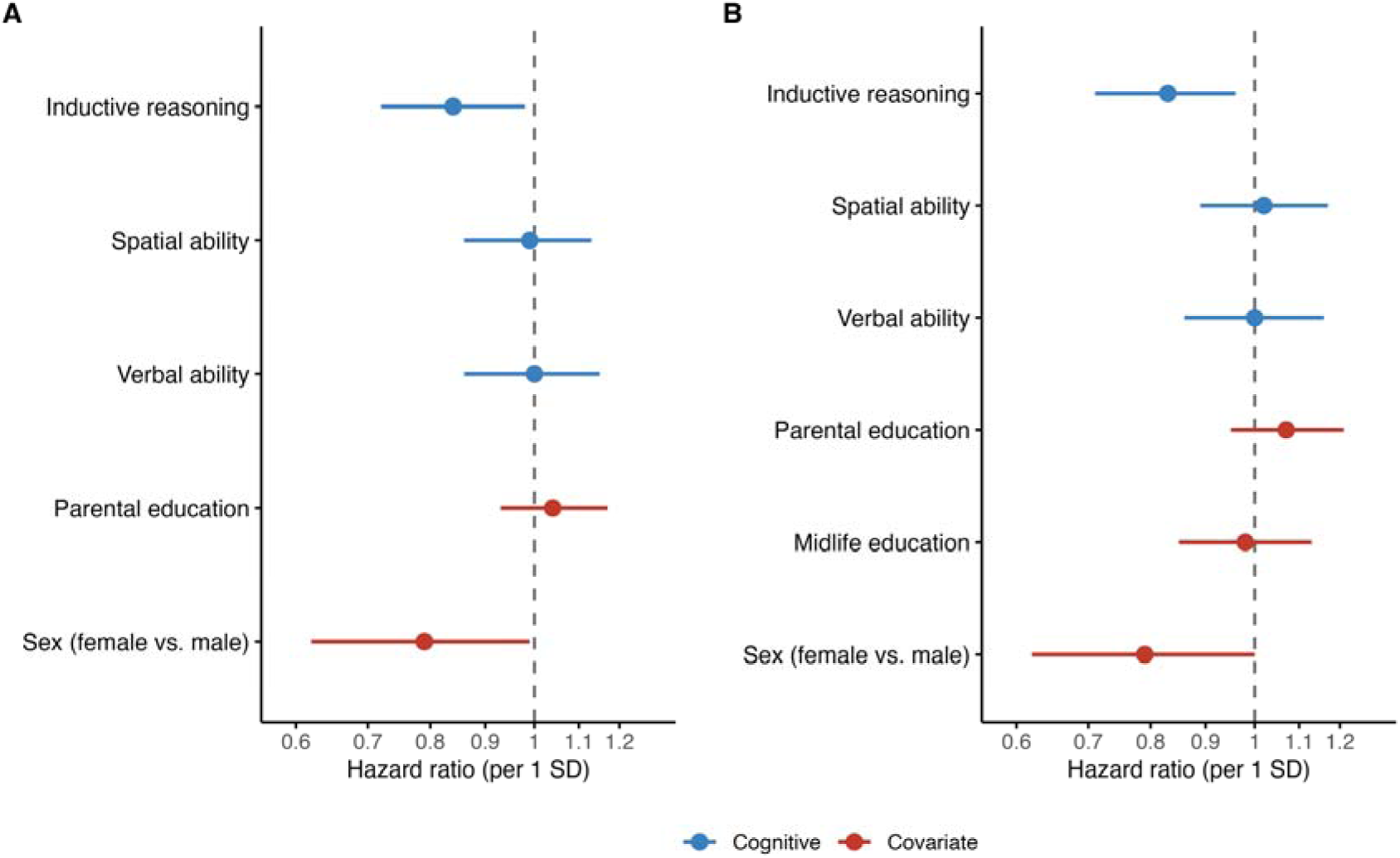
Childhood cognitive scores, education, sex and dementia risk. Forest plots showing hazard ratios and 95% confidence intervals for dementia associated with standardized childhood cognitive test scores (per 1 SD), education and sex (female vs. male) in the 1948 cohort. **A) Parental education** is entered as a covariate in the model (n = 10, 539; 287 dementia cases). Higher inductive reasoning scores at age 13 were associated with lower hazard of dementia, and being female was associated with lower hazard, whereas verbal, spatial, and parental education effects were not statistically significant (Table S5). **B) Parental education and participants’ own educational midlife attainment** recorded in the 1990 Census are entered as covariates (n = 9, 640; 280 dementia cases). Higher childhood inductive reasoning remained associated with lower dementia risk after adjustment for adult educational attainment, whereas verbal and spatial abilities were not. Sex also remained associated, with women having lower dementia risk than men (Table S6).

As a post-hoc follow-up test, each cognitive domain was next modelled individually. Only inductive reasoning remained significantly associated with dementia risk (HR 0·83, 95% CI 0·74–0·94), with spatial and verbal abilities remaining non-significant (Supplementary results), confirming that the domain-specific pattern was not an artefact of joint modelling.

Sensitivity analyses correcting for minor subtest reliability differences yielded virtually identical results. Competing risk from death was formally addressed using Fine–Gray sub-distribution hazard models, yielding results consistent with the primary Cox models. Recorded emigration does not necessarily equal loss to follow-up in Swedish registry data, but results were also unchanged when excluding 262 participants with recorded emigration and no subsequent CDR death (Supplementary Results).

We added and tested an interaction between sex and inductive reasoning, as differential sex associations of childhood and adolescent cognition with dementia risk have been suggested by previous reports,(4, 10) however, this was not significant (HR = 0.94, 95% CI: 0.74–1.19).

### Adjustment for adult educational attainment

We additionally adjusted the full model for participants’ own midlife educational attainment (Figure 1B, Table S6). 9640 participants had complete data, including 280 dementia cases. The association between childhood inductive reasoning and dementia remained virtually unchanged (HR per SD = 0.83, 95% CI: 0.71–0.96), and other associations also remained similar. Adult educational attainment was not independently associated with dementia risk.

### Somatic morbidity, cardiovascular and cardiometabolic disease analyses

During follow-up 5,183 participants in the 1948 cohort experienced at least one major somatic morbidity condition. In Cox models with covariate specification identical to the primary dementia models, higher childhood inductive reasoning associated with lower risk of major somatic morbidity (HR per SD, 0.91; 95% CI, 0.88–0.94), which strongly associated with dementia risk (HR, 1.63; 95% CI, 1.28–2.07). However, inclusion of somatic morbidity in the dementia model resulted in only modest attenuation of the association between childhood inductive reasoning and dementia (HR, 0.85; 95% CI, 0.73–0.99), and associations were upheld when also including midlife education as a covariate (Tables S8-S10).

Higher childhood inductive reasoning associated with lower risk of each cardiovascular outcome alone and when combined into a composite (HR, 0.88; 95% CI, 0.83–0.92), as well as with diabetes (HR, 0.74, 95% CI, 0.64–0.86). Each cardiovascular condition was also associated with increased dementia risk alone and as a composite (HR, 2.00; 95% CI, 1.53–2.63). Diabetes also associated strongly with dementia risk (HR, 2.74; 95% CI, 1.53–4.88). These associations were upheld when including also midlife education as a covariate. However, inclusion of cardiovascular (Figure 2) and cardiometabolic conditions in dementia models only modestly attenuated the association between childhood inductive reasoning and dementia (HR ≈ 0.84-0.86 across models). While higher midlife education related to lower risk of diseases, it was not a significant predictor of dementia when entered together with the somatic diseases and cognitive tests. Relations between inductive reasoning, other somatic disease outcomes and dementia were modestly attenuated and remained significant when controlling for midlife education. Although cardiovascular and cardiometabolic conditions were associated with increased dementia risk, overlap with dementia cases was incomplete, with a substantial proportion occurring without recorded cardiovascular or cardiometabolic disease: among 287 dementia cases, 168 had no recorded cardiovascular disease or diabetes prior to or at diagnosis, whereas 103 had CVD only, 8 had diabetes only, and 8 had both, consistent with the minimal attenuation of the inductive reasoning–dementia association after adjustment. For a summary, see Table 2, and for details of all analyses see Tables S11-S24.

**Figure 2.**
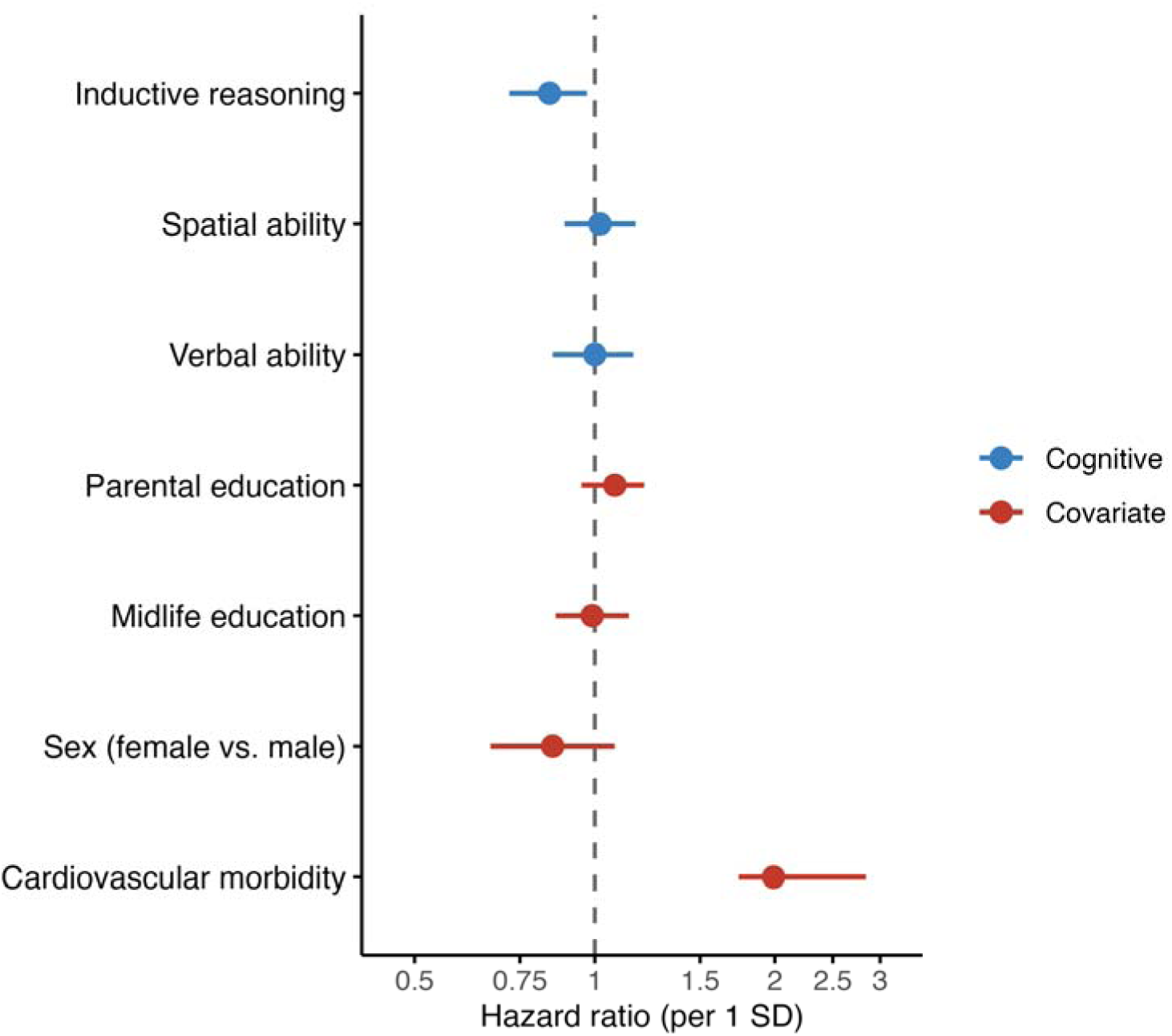
Childhood cognitive scores, parental and adult educational attainment, cardiovascular morbidity, sex, and dementia risk. Forest plot showing hazard ratios (HRs) and 95% confidence intervals for dementia associated with standardized childhood cognitive test scores (per 1 SD), parental education, participants’ own educational midlife attainment recorded in the 1990 Census, cardiovascular morbidity, and sex in the 1948 cohort. Cox proportional hazards models used age as the underlying time scale. Cardiovascular morbidity was modeled as a time-varying covariate based on first inpatient diagnosis of myocardial infarction, heart failure, or cerebrovascular disease. Higher childhood inductive reasoning was associated with lower dementia risk; cardiovascular morbidity was strongly associated with increased dementia risk. The horizontal axis is displayed on a logarithmic scale. For details, see Supplement, Table S20.

**Table 2.** Association between childhood inductive reasoning and dementia across all model specifications (n = 10,539, 287 dementia cases) Hazard ratios (HR) and 95% confidence intervals represent the association between childhood inductive reasoning (per SD) and dementia risk, adjusted for parental education and sex in all models. The primary model establishes inductive reasoning as a predictor of dementia risk. Subsequent models additionally adjust for clinically manifest major somatic morbidity, cardiovascular disease, and diabetes as time-varying covariates, entered separately, to assess whether the association between inductive reasoning and dementia persisted after accounting for these conditions. Morbidity covariates were modelled as time-varying, switching from 0 to 1 at the age of first diagnosis. For models including midlife education, n = 9,640.

| **Model** | **HR** | **95% CI** |
| --- | --- | --- |
| Primary (no morbidity covariate) | 0.84 | 0.72–0.98 |
| Somatic morbidity as time-varying covariate | 0.85 | 0.73–0.99 |
| Composite CVD as time-varying covariate | 0.85 | 0.73–0.99 |
| Cerebrovascular disease as time-varying covariate | 0.85 | 0.73–0.98 |
| Myocardial infarction as time-varying covariate | 0.84 | 0.73–0.98 |
| Heart failure as time-varying covariate | 0.85 | 0.73–0.98 |
| Diabetes as time-varying covariate | 0.85 | 0.73–0.98 |
| Composite CVD + midlife education | 0.84 | 0.72–0.97 |
| Somatic morbidity + midlife education | 0.84 | 0.72–0.97 |

### Analysis in an expanded cohort

To evaluate robustness of results, we added the UGU 1953 birth cohort, yielding a total of 19,919 individuals (10,100 males) and 367 dementia cases (1.84%). This cohort was tested at age 13 years in 1966 and thus had 5 years less follow-up time and lower age than the 1948 cohort (see Supplement). We were thus not powered to analyze this cohort separately, and data on the equivalent subtest in this cohort was only used to assess stability of the finding form the 1948 cohort. Adjusted for sex, inductive reasoning associated with lower dementia risk with comparable effect size (HR = 0.85, 95% CI: 0.77–0.95, p = 0.003). To assess potential non-linear associations (5), we fitted a generalized additive model with a penalized spline. The relationship was essentially linear (Figure 3). A formal comparison between the GAM and a simple logistic regression model revealed no improvement in fit for the non-linear specification (Supplementary Results).

**Figure 3.**
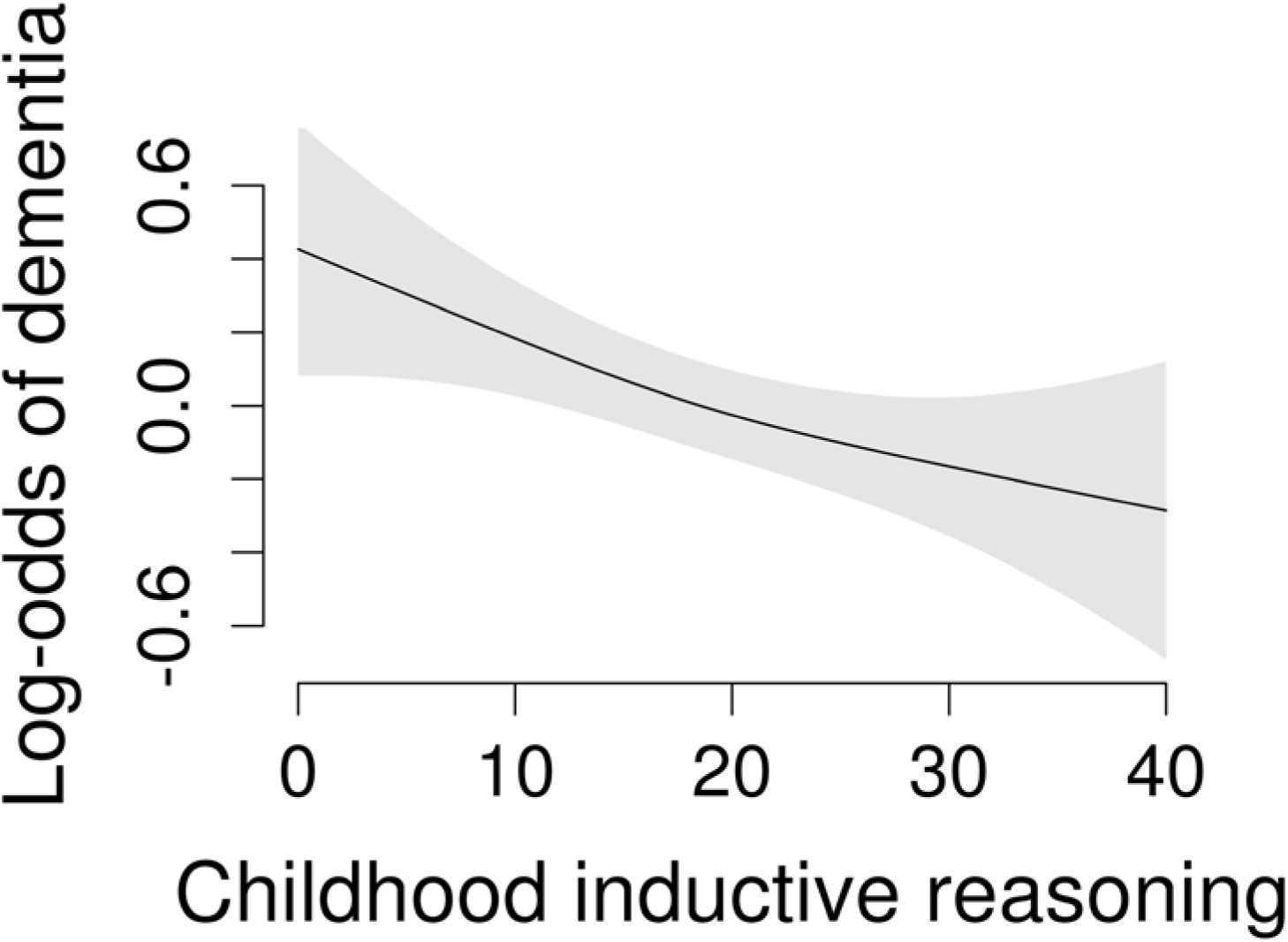
Inductive reasoning and dementia risk in the expanded cohort. Generalized additive model smooth showing the association between inductive reasoning at age 13 and later-life dementia risk in the expanded cohort (1948 and 1953 birth cohorts; n = 19,919). The solid line indicates the estimated effect on the log-odds scale, and the shaded region shows the 95% confidence interval of the smooth line. The relationship is essentially linear, with higher childhood reasoning scores associated with lower dementia risk.

## Discussion

Our objectives were to test whether specific cognitive domains are uniquely predictive of dementia independently of pre-test educational differences, and when adjusting for other major diseases. In this population-based study with more than six decades of follow-up, we found that childhood cognitive ability is related to dementia risk in a domain-specific manner. Higher inductive reasoning performance at age 13 was associated with reduced risk of dementia in late life, whereas verbal and spatial ability scores were not independently associated. For every additional raw point scored on the number series test in childhood, dementia risk was approximately 2% lower, corresponding to an estimated difference in risk of roughly 30% across the observed ability range. These associations were evident in a cohort with minimal selection bias when controlling for parental and own midlife education differences and sex, as well as incident major somatic, cardiovascular and cardiometabolic conditions. Together with work linking higher early-life cognition to lower dementia risk (2–5), our findings suggest that individual differences in inductive reasoning before educational attainment diverges may represent one component of life-course resilience related to maintaining cognitive function above clinical thresholds (26), not substantially attenuated by adjustment for education and clinically manifest somatic disease.

### Developmental timing

Most registry studies linking early cognition to dementia have assessed ability in late adolescence or early adulthood, typically at military conscription (5, 8, 9, 27), when test performance is already shaped by schooling and related social factors (28). In contrast, our assessments were conducted at age 13, when educational variation was limited, reducing potential confounding by completed schooling.

The Scottish Mental Surveys showed that global cognitive ability at age 11 predicted dementia risk, particularly among women (2–4). Extending this work, we found that inductive reasoning specifically at age 13 associated with dementia risk over six decades in a population-based cohort. Unlike the Scottish studies, we observed no sex interaction, and this may relate to societal differences or historic changes between cohorts. Lower dementia risk among women corresponds with national patterns in these birth cohorts (29). Taken together, the findings are consistent with a life-course model where aspects of resilience and vulnerability are established early even if shaped by subsequent accumulated exposures (26, 30, 31). Inductive reasoning in childhood may index neurodevelopmental characteristics that precede and are not fully captured by formal education, since the observed relationship was not explained by subsequent educational attainment, in turn not significantly associated with dementia risk when childhood cognitive abilities were included in the model.

Our findings indicate that the association between childhood inductive reasoning and dementia is unlikely to merely reflect a nonspecific gradient in overall health or cardiovascular and cardiometabolic disease alone, consistent with the incomplete overlap of these conditions with dementia cases observed in the cohort. One hypothesis is that cognitive ability relates to later health because it is an index of general bodily “system integrity” (32). This is not undermined by the current findings, but partly different factors may additionally be at play in determining different types of health outcomes. Cognitive ability overlaps genetically with dementia, cardiovascular, cardiometabolic and other diseases (33, 34), and genetic factors may also underlie the currently observed associations. Although inductive reasoning, consistent with prior evidence linking early cognitive differences to later health outcomes (1, 3, 11), was also associated with incident CVD and diabetes, and these conditions are strongly associated with dementia (13), adjustment for clinically manifest CVD and diabetes only modestly attenuated the association with dementia. These results suggest that the life-course association between childhood inductive reasoning and dementia is not primarily explained by overt cardiovascular or metabolic disease, even if inductive reasoning shows domain-specific associations with these conditions.

### Domain-specific vulnerability

Inductive reasoning was the only childhood cognitive domain independently associated with later-life dementia risk. This finding is novel, yet aligns with evidence that fluid-type processes are more sensitive to early neurodegenerative change than crystallized verbal abilities (35) and that fluid components load strongly on general intelligence (g) (36), central to other cohort studies (2–5). Reasoning tasks engage frontoparietal systems implicated in age-related decline and Alzheimer disease pathology, potentially indexing early neurodevelopmental robustness rather than accumulated knowledge systems (37, 38). Reasoning also tends to show strong associations with educational attainment, occupational complexity and lifelong cognitive engagement, relating to dementia risk (39, 40). As such, results align with evidence from epidemiology and cognitive neuroscience.

Although adjustment for cardiovascular and cardiometabolic disease only modestly attenuated the association with dementia, childhood inductive reasoning was also specifically associated with such conditions. Cognitive ability when young has been robustly associated with later cardiovascular and cardiometabolic disease (12, 14, 16). While downstream factors such as education often attenuate child ability-later health relationships, they also tend to persist when adjusted for such (12, 14, 16), suggesting broader developmental patterns. For instance, family SES and pre- and perinatal conditions relate to childhood cognitive function, CVD and dementia (17, 41–45). However, even in studies adjusting for such factors, associations between childhood cognition and CVD remain(15, 16), and unmeasured downstream risk factors, e.g. via health behaviors and risks are also likely.

Since inductive reasoning reflects core fluid cognitive processes shaped early in life and linked to lifelong learning capacity, the findings may have policy implications. Quality of environments may influence whether the cognitive potential of children is reached. Increases in cognitive scores between cohorts over time, the Flynn effect, tend to be greater for fluid tests, indicating influence by societal change (46). Interestingly, the current cohort grew up in a time when one saw such population level rises in cognitive function, now coinciding with currently decreasing dementia incidence (29, 46). Increased mandatory schooling in adolescence has been associated with increase in intelligence test performance (28). However, while crystallized scores increase with schooling, fluid scores appear not to (47). Correspondingly, extending primary education has shown negligible effects on dementia risk (48). This, coupled with recent evidence that the association of education and dementia is largely accounted for by cognitive ability (5), dampen hopes of prevention by increasing educational amount. That does however not imply that neurocognitive developmental robustness to disease cannot be boosted. Here, insight in what cognitive subdomain matters most, is highly relevant.

Inductive reasoning is not a fixed trait in childhood (49–51).Despite little transfer of targeted training effects on general cognitive ability (52, 53), inductive reasoning can persistently be improved for children through training (51). Policies that enhance child environments may strengthen neurodevelopmental foundations that later buffer against falling below a cognitive functional threshold (26). This supports a shift toward a public health framework recognizing early cognitive development as a critical factor for later-life dementia risk. In context, a 4-raw point difference in test score at age 13 associated with a magnitude of dementia risk comparable to established midlife risk factors identified by the Lancet commission on dementia prevention (13).

### Limitations

The nationally representative primary school cohort, combined with population-wide ascertainment of dementia from the NPR-IPR and CDR, reduces selection bias related to socioeconomic status and healthcare access. Because hospitalizations and deaths are comprehensively registered, outcome definitions are likely less influenced by help-seeking behavior than outpatient or insurance-based records(20). However, such registry-based dementia diagnoses have imperfect sensitivity and likely capture cases later in the disease course (21). By end of observation (2025), the primary cohort had reached approximately age□77. As peak dementia incidence occurs in the ninth decade, continued follow-up will yield additional events. Associations may differ in outpatient settings and older cohorts. We adjusted for parental education, midlife educational attainment, and major somatic, cardiovascular, and cardiometabolic disease, but did not measure prenatal and early life health conditions or subsequent occupation and midlife health behaviors, and still cannot determine whether childhood reasoning influences dementia risk directly, through other upstream or downstream exposures (1, 11, 31, 54–57), or represents a marker of other conditions of relevance for both. We did not examine dementia subtypes, nor include all potentially relevant cognitive domains (e.g., memory). Residual confounding by familial, genetic (33, 34), or other health factors cannot be excluded. A sex-interaction term was tested and was not significant. Analyses were not stratified by sex; sex was included as a covariate in all models, but sex-specific effect estimates were not the primary focus. The UGU cohort is drawn from a Swedish population-representative birth cohort, and data on ethnicity specifically were not collected. Generalizability to more diverse populations is uncertain.

## Conclusions

In this nationally representative Swedish cohort followed for more than six decades, higher inductive reasoning at age 13 was associated with lower dementia risk and was not substantially attenuated by adjustment for education or clinically manifest somatic disease. While causality cannot be inferred from these findings, they extend previous evidence on associations of early life cognitive function and later disease. Collectively, this supports a life-course perspective in which specific early neurodevelopmental differences relate to later dementia vulnerability, alongside established vascular and metabolic risk factors.

## Data Availability

Individual-level data from the Evaluation Through Follow-up (UGU) study and linked Swedish national health registers are not publicly available because they are subject to Swedish and EU data protection legislation and ethical approvals that restrict data sharing. Researchers may apply for access to the UGU data through the University of Gothenburg and, where applicable, to linked Swedish national register data through the relevant Swedish authorities, subject to ethical approval and data protection agreements. Information about the application process for UGU data is available at: https://www.gu.se/en/evaluation-through-follow-up/disclosure-of-ugu-data. Statistical code developed by the authors and used for analyses in the current study specifically is shared here: https://github.com/UGU-LIFE/child-cognition-cardio-health-dementia/tree/main

https://github.com/UGU-LIFE/child-cognition-cardio-health-dementia/tree/main

## List of abbreviations

CCI: Charlson Comorbidity Index
CDR: Cause of Death Registry
CI: Confidence interval
CVD: Cardiovascular Disease
HR: Hazard ratios
IQR: interquartile range
M: Mean
NPR-IPR: National Patient Registry - Inpatient Registry
SD: Standard deviation
SES: Socio-economic status
VIFs: variance inflation factors

## Declarations

### Ethics approval and consent to participate

Approval for the study was given by the Swedish Ethical Review Authority (case number 2024-06028). Consent was not necessary. Individuals in the study population were given the opportunity to opt-out of registry linkage (see below).

### Consent for publication

Not applicable

### Authoŕs Contributions

KBW and ML (M. Lövdén) conceived and designed the study. KBW conducted data analysis and wrote the manuscript. All authors took part in concept and design, undertook critical revision of the manuscript for important intellectual content, and are responsible for the decision to submit the manuscript. KBW, ML and AS directly accessed and verified the underlying data reported in the manuscript. AS ran an independent test of dementia code extraction in Python to verify equivalence of registry extraction across software.

## Acknowledgements

We thank all participants in the UGU 1948 and 1953 cohorts, Lucas Fischer Madsen for database management, and Ole J. Røgeberg and Anders M. Fjell for comments to the manuscript. Kristine B. Walhovd, affiliated with the University of Gothenburg, Sweden, and University of Oslo, Norway, conducted and is responsible for the data analysis. Kristine B. Walhovd had full access to all the data in the study and takes responsibility for the integrity of the data and the accuracy of the data analysis. Walhovd used ChatGPT version 5.2 (OpenAI, San Francisco, CA, USA) and Claude Sonnet 4.6 (Anthropic, San Francisco, CA) to help shorten the abstract, and to help debug part of the code written by Walhovd. Walhovd takes responsibility for the integrity of the content thus revised by this tool and revised all contents herself.

