## Supplement for "Childhood inductive reasoning, cardiovascular and cardiometabolic morbidity and dementia risk – a population representative cohort study across six decades"

for

**Content:**	p

**Supplementary Methods**	3

Figure S1 Flow chart of 1948 sampling frame, exclusions and analytic sample	3

Table S1. Characteristics of the 1948 birth cohort by missing midlife education	4

Cognitive tests – reliabilities and subtest correlations	5

Figure S2 Distributions of childhood cognitive scores	5

Education measures and distributions	6

Details of dementia and other disease outcome coding	6

Definition of somatic major morbidity	7

Definition of cardiovascular disease and diabetes	8

Table S2. Characteristics of the 1948 birth cohort by somatic morbidity status	10

Table S3. Characteristics of the 1948 birth cohort by CVD status	11

Table S4. Characteristics of the 1948 birth cohort by diabetes status	12

Supplementary statistical analyses	13

Statistical analyses of diseases other than dementia	14

**Supplementary Results**	15

Table S5 Child cognition and dementia, main model	15

Table S6 Child cognition and dementia, main model, education-adjusted	15

Sensitivity Analyses	16

*Post-hoc tests of isolated subtests to address potential partialling problem* 16

*Subtest reliability correction*	16

*Sensitivity to birth month*	17

*Analysis of possible loss to follow-up by migration*	17

*Analysis of competing risk*	17

Table S7 Child cognition and major somatic morbidity	18

Table S8 Child cognition and major somatic morbidity, education-adjusted	18

Table S9 Dementia model adjusted for major somatic morbidity	19

Table S10 Dementia model adjusted for major somatic morbidity and education	19

Table S11 Child cognition and cerebrovascular disease	20

Table S12 Dementia model adjusted for cerebrovascular disease	20

Table S13 Child cognition and myocardial infarction	21

Table S14 Dementia model adjusted for myocardial infarction	21 Table S15 Child cognition and congestive heart failure	21

Table S16 Dementia model adjusted for congestive heart failure	21

Table S17 Child cognition and any cardiovascular disease	23

Table S18 Child cognition and any cardiovascular disease, education-adjusted	23

Table S19 Dementia model adjusted for any cardiovascular disease	24

Table S20 Dementia model adjusted for any cardiovascular disease, education	24

Table S21 Child cognition and diabetes	25

Table S22 Child cognition and diabetes, education-adjusted	25

Table S23 Dementia model adjusted for diabetes	26

Table S24 Dementia model adjusted for diabetes and education	26

Details on analysis results in the expanded cohort	27

**References**	29

**Supplementary Methods**

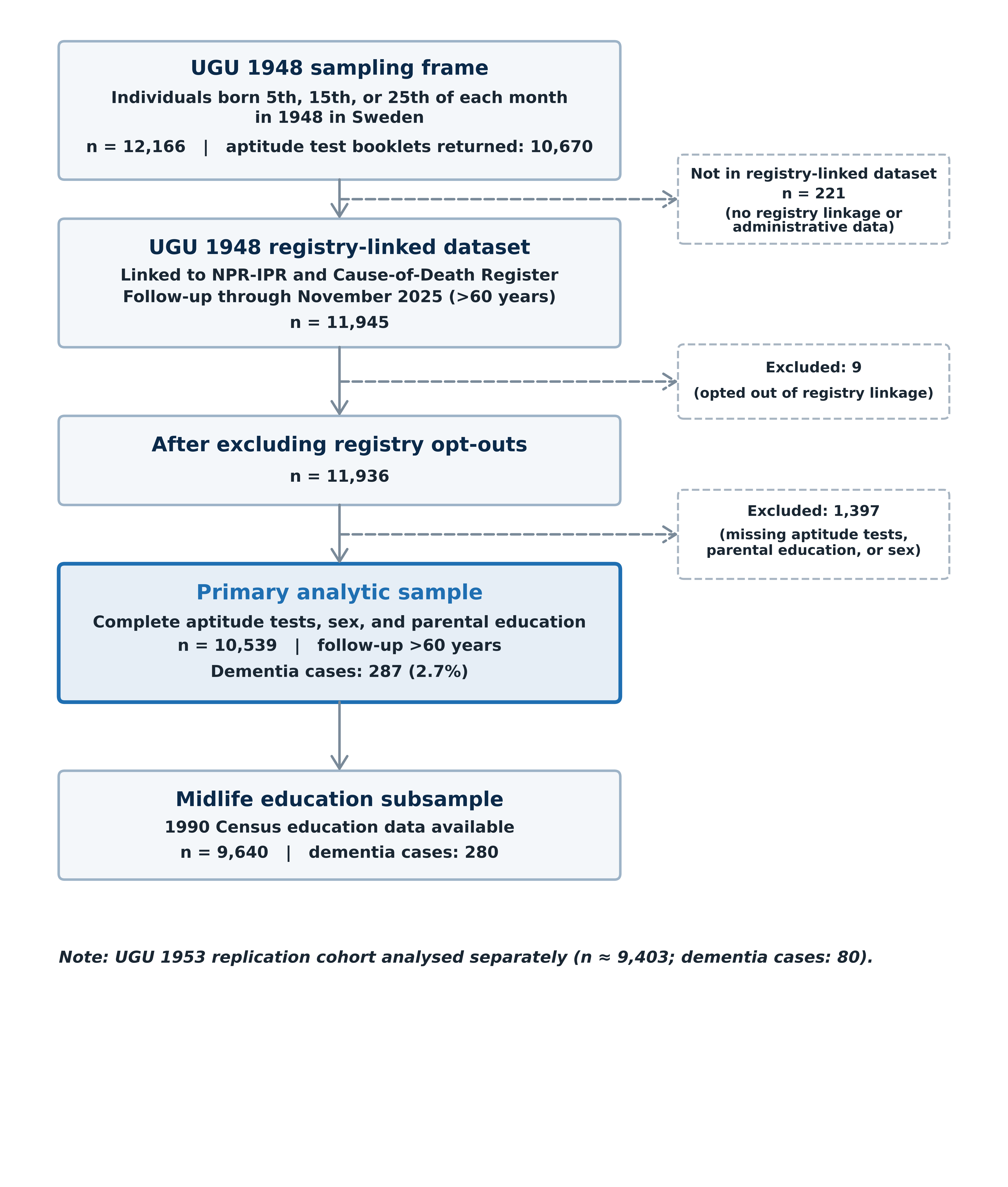

***Figure S1 Flow chart of 1948 sampling frame, exclusions and analytic sample*** The figure shows how the analytic sample relates to the maximum possible population. Protocol descriptions can be found here: <https://www.gu.se/en/evaluation-through-follow-up> with further links to details for the 1948 and 1953 cohorts used.(1)

**Table S1. Characteristics of the 1948 Birth Cohort According to Availability of Midlife Education Data**

Values are presented as n (%) for categorical variables, mean (SD) for continuous cognitive and education measures, and median (interquartile range [IQR]) for age variables. Parental education was coded on an ascending scale from 1 to 4. Midlife education obtained from the 1990 Swedish Census was missing for 899 of 10539 participants in the 1948 cohort, including 7 dementia cases.

| **Characteristic** | **Non-missing (n=9,640)** | **Missing**  **(n=899)** |
| --- | --- | --- |
| Female, n (%) | 4780 (49.6%) | 387 (43.0%) |
| Inductive reasoning, mean (SD) | 19.34 (7.95) | 19.27 (8.05) |
| Spatial ability, mean (SD) | 21.01 (7.24) | 21.48 (7.27) |
| Verbal ability, mean (SD) | 22.24 (6.98) | 23.04 (7.33) |
| Parental education, mean (SD) | 1.22 (0.53) | 1.34 (0.65) |
| Age at dementia diagnosis, median (IQR), y | 73.62 (5.66) | 69.46 (5.52) |

*Cognitive tests – reliabilities and subtest correlations*

Test scores have been reported to have acceptable reliabilities: antonyms: .87; metal folding: .88; number series: .92,(2) and correlated within expected range: antonyms-number series: .56, antonyms-metal folding: .42, metal folding-number series: .48. While subtest reliabilities are all acceptable with only slight differences, we conducted a reliability sensitivity analysis adjusting for differences in reliability, since the study targets domain-specificity (see p. 16, Supplementary results, Sensitivity analyses). The raw subtest score distributions are shown in Figure S2 below.

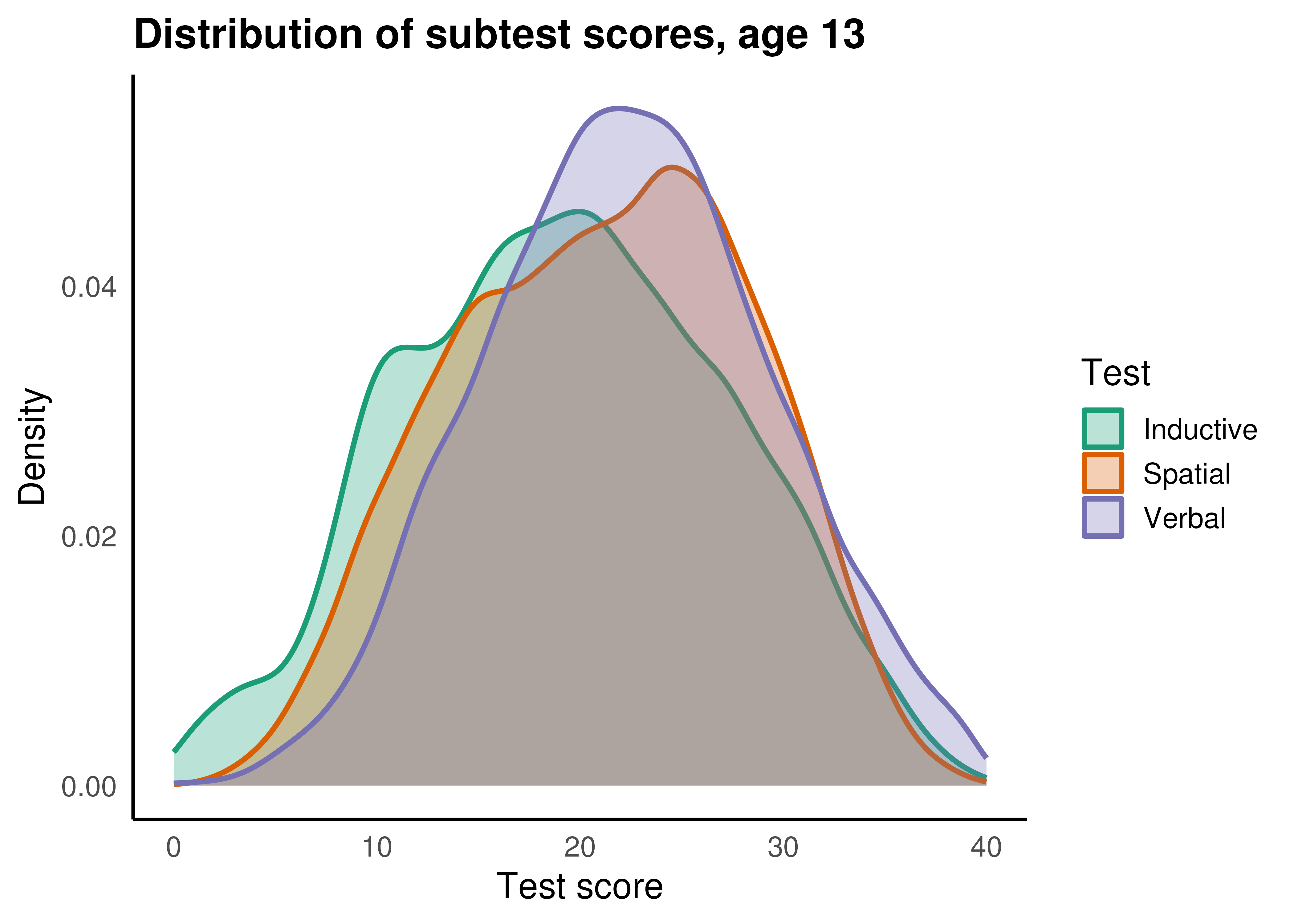

***Figure S2. Distributions of childhood cognitive scores.***

Kernel density distributions of the three cognitive tests administered at age 13: inductive reasoning, spatial ability, and verbal ability. The curves represent smoothed density estimates, illustrating differences in central tendency and spread across subtests.

*Education measures and distributions*

Parental education was recorded as a school administrative variable at the time of testing and coded on an ascending scale from 1 to 4. The mean of paternal and maternal education, or either parent´s education if one was missing (N = 355 fathers, 59 mothers), was used in analyses (M = 1.227, SD = 0.543). Of those with reported education, 84,6% of fathers and 85.8% of mothers had only *elementary school*, 6.6% and 11.0% had *secondary school*, 5.3% and 2.4% had *upper secondary school or equivalent*, and only 3.5% and 0.7% held a *diploma,* respectively. Participants’ individual educational attainment was recorded in the 1990 Census (Folk och Bostadsräkningen) when individuals were ~42 years of age. This midlife education measure was available for 9640 persons of those in the 1948 cohort who had also the other measures. Midlife educational attainment recorded in the 1990 Census was classified on a 7-level scale reflecting the highest completed level of formal education. Among men, 19.0% had pre-upper secondary education of ≤9 years and 16.4% had exactly 9 years of pre-upper secondary education, compared with 15.0% and 19.5%, respectively, among women. Upper secondary education of up to 2 years was completed by 18.7% of men and 26.1% of women, whereas longer upper secondary education (>2 and ≤3 years) was more common among men (17.9%) than women (8.3%). Short post-secondary education (<3 years, including 4-year upper secondary programs) was completed by 10.1% of men and 15.6% of women, while long post-secondary education (≥3 years) was similarly distributed (16.7% of men and 15.4% of women). Doctoral education was rare in both sexes (1.1% of men and 0.3% of women).

**Details of dementia and other disease outcome coding**

Dementia diagnoses were identified in the Swedish National Patient Register – In-patient Register (NPR-IPR) and Cause of Death Register (CDR) using International Classification of Diseases codes (ICD), ninth and tenth revision. Of the 287 dementia cases in the analytic sample, 43 were recorded only in CDR, 83 in both CDR and NPR-IPR, the remaining 161 only in NPR-IPR. Major somatic, cardiovascular and cardiometabolic diseases were identified in NPR-IPR across the lifespan using ICD7-10 diagnoses. Data were retrieved from registries until November 3^rd^ 2025. Very few of the sampled individuals would be eligible for a dementia diagnosis in ICD9, used in Sweden from 1985 until 1997/98. For ICD9, codes 290 and 331, Swedish subcodes A, B, and C were sampled. For ICD 10, we captured all codes starting with F00, F01, F02, F03, and G30, and additionally codes G310, G318 and G319. A person was classified as having dementia if any available diagnosis for hospitalization or death, main or supporting, contained a relevant code. Time of diagnosis was set to earliest date of hospitalization or time of death.

*Definition of major somatic morbidity*

To examine whether associations between childhood cognitive performance and dementia risk were attenuated by adjustment for medical morbidity, we conducted parallel time-to-event analyses using diagnostic categories from an index of overall somatic major morbidity based on the Charlson Comorbidity Index (CCI)(3, 4). The CCI is a disease index, listing ICD codes from the following diseases beyond dementia: myocardial infarction, congestive heart failure, peripheral vascular disease, cerebrovascular disease, pulmonary diseases, rheumatic disease, hemiplegia, diabetes, chronic kidney disease, liver disease, peptic ulcer disease, cancer and HIV/AIDS. Diagnoses were identified from the NPR-IPR using a validated Swedish adaptation of the Charlson algorithm spanning ICD-7 through ICD-10 classification systems(4, 5) (based on updated version from Dec 17^th^ 2025 <https://github.com/bjoroeKI/Charlson-comorbidity-index-revisited/blob/main/Charlson_R>). For each participant, we identified the earliest recorded diagnosis corresponding to any Charlson condition, and in keeping with their recommendation, we used diagnoses at the date of discharge from hospital.

Because dementia is included in the Charlson classification, dementia diagnoses were excluded from the composite outcome to avoid overlap with the primary endpoint. Only diagnoses set prior to, or concurrent with, the occurrence of a first dementia diagnosis if ever, were included as covariates in the models predicting dementia, as our goal was to study to what extent somatic morbidity, and CVD or diabetes specifically, preceded dementia onset.

*Definition of Cardiovascular Disease and Diabetes*

Cardiovascular and cardiometabolic conditions were likewise defined as separate categories for analyses using inpatient diagnoses from the NPR-IPR ICD-7-10 classification systems. For each condition, the earliest recorded inpatient diagnosis at date of hospital discharge was identified and used as the event date. Individuals were classified as exposed from the date of first qualifying diagnosis onward in time-varying models.

*Myocardial infarction* was defined using the following ICD codes: ICD-7: 420.1; ICD-8: 410, 411, 412.01, 412.91; ICD-9: 410, 412; ICD-10: I21, I22, I252. *Congestive heart failure* was defined as: ICD-7: 422.21, 422.22, 434.1, 434.2; ICD-8: 425.08, 425.09, 427.0, 427.1, 428; ICD-9: 402A, 402B, 402X, 404A, 404B, 404X, 425E, 425F, 425H, 425W, 425X, 428; ICD-10: I110, I130, I132, I255, I420, I426, I427, I428, I429, I43, I50. *Cerebrovascular disease* was defined as: ICD-7: 330–334; ICD-8: 430–438; ICD-9: 430–438; ICD-10: G45, I60–I64, I67, I69. A *composite cardiovascular disease variable* was defined as the first occurrence of myocardial infarction, congestive heart failure, or cerebrovascular disease.

*Diabetes* was categorized as with or without chronic complications, based on inpatient diagnoses. *Diabetes without chronic complications* was defined as: ICD-7: 260.09; ICD-8: 250.00, 250.07, 250.08; ICD-9: 250A, 250B, 250C; ICD-10: E100, E101, E106, E109, E110, E111, E119, E120, E121, E129, E130, E131, E139, E140, E141, E149. *Diabetes with chronic complications* was defined as ICD-7: 260.2, 260.21, 260.29, 260.3, 260.4, 260.49, 260.99; ICD-8: 250.01–250.05; ICD-9: 250D, 250E, 250F, 250G; ICD-10: E102–E107, E112–E117, E122–E127, E132–E137, E142–E147. If both categories were recorded, individuals were classified as having diabetes with chronic complications.

**Table S2. Characteristics of the 1948 Birth Cohort According to Presence of Major Somatic Morbidity**

Values are presented as n (%) for categorical variables, mean (SD) for continuous cognitive and education measures, and median (interquartile range [IQR]) for age variables. Major somatic morbidity was defined using the time-based Charlson Comorbidity Index derived from nationwide register data on hospitalizations (NPR-IPR).

| **Characteristic** | **No somatic morbidity (n=5,356)** | **Somatic morbidity (n=5,183)** |
| --- | --- | --- |
| Female, n (%) | 2801 (52.3%) | 2366 (45.6%) |
| Inductive reasoning, mean (SD) | 19.87 (7.91) | 18.78 (7.97) |
| Spatial ability, mean (SD) | 21.27 (7.23) | 20.83 (7.25) |
| Verbal ability, mean (SD) | 22.66 (6.98) | 21.95 (7.03) |
| Parental education, mean (SD) | 1.26 (0.58) | 1.20 (0.50) |
| Midlife education, mean (SD) | 3.51 (1.72) | 3.25 (1.68) |
| Age at dementia diagnosis, median (IQR), y | 73.34 (6.29) | 73.62 (5.78) |

### **Table S3. Characteristics of the 1948 birth cohort according to presence of any of the cardiovascular disorders**

Values are presented as n (%) for categorical variables, mean (SD) for continuous cognitive and education measures, and median (interquartile range [IQR]) for age variables. Composite cardiovascular disorder was defined using nationwide register data on hospitalizations and included any of the recorded cardiovascular conditons (cerebrovascular disorder, myocardial infarction, congestive heart failure) during follow-up.

| **Characteristic** | **No cardiovascular disorder (n=8,172)** | **Cardiovascular disorder (n=2,367)** |
| --- | --- | --- |
| Female, n (%) | 4270 (52.3%) | 897 (37.9%) |
| Inductive reasoning, mean (SD) | 19.55 (7.93) | 18.59 (8.02) |
| Spatial ability, mean (SD) | 21.07 (7.25) | 21.01 (7.24) |
| Verbal ability, mean (SD) | 22.45 (6.99) | 21.81 (7.08) |
| Parental education, mean (SD) | 1.24 (0.55) | 1.20 (0.52) |
| Midlife education, mean (SD) | 3.42 (1.71) | 3.23 (1.70) |
| Age at dementia diagnosis, median (IQR), y | 73.36 (5.58) | 73.75 (6.64) |

**Table S4. Characteristics of the 1948 birth cohort according to presence of Diabetes**

Values are presented as n (%) for categorical variables, mean (SD) for continuous cognitive and education measures, and median (interquartile range [IQR]) for age variables. Diabetes was defined using nationwide register data on hospitalizations and included any recorded diagnosis during follow-up.

| **Characteristic** | **No diabetes (n=10,231)** | **Diabetes (n=308)** |
| --- | --- | --- |
| Female, n (%) | 5057 (49.4%) | 110 (35.7%) |
| Inductive reasoning, mean (SD) | 19.41 (7.94) | 16.87 (8.21) |
| Spatial ability, mean (SD) | 21.09 (7.23) | 19.98 (7.64) |
| Verbal ability, mean (SD) | 22.35 (7.01) | 20.81 (6.95) |
| Parental education, mean (SD) | 1.23 (0.55) | 1.13 (0.37) |
| Midlife education, mean (SD) | 3.39 (1.71) | 2.92 (1.59) |
| Age at dementia diagnosis, median (IQR), y | 73.60 (5.82) | 72.68 (5.95) |

**Supplementary statistical analyses**

Analyses were performed in R v4.3.3. We estimated hazard ratios (HRs) and 95% confidence intervals using Cox proportional hazards regression with age as the underlying timescale, to predict dementia (0 = no dementia, 1 = dementia):

*Surv(start_age, end_age, dementia) ~ inductive reasoning + spatial ability + verbal ability + parental education + sex.*

All continuous predictors were standardized, and sex was coded as 0 = male, 1 = female. Models were fitted using the coxph function from the *survival* package in R. Proportional hazards assumptions were evaluated using Schoenfeld residuals and the global test from cox.zph() (survival package). No evidence of violation was found for any predictor or globally (global χ² = 3.75, df = 5, p = 0.59). To assess the joint contribution of the three cognitive measures, we compared a reduced model including only sex and parental education with a full model including all cognitive predictors using a likelihood ratio test. Participants’ educational attainment recorded in the 1990 Census was included in secondary analyses to examine whether associations were robust to adjustment for midlife education. We next conducted parallel time-to-event analyses where cardiovascular conditions, including cerebrovascular disease, myocardial infarction, and heart failure, as well as diabetes were modeled as independent outcomes predicted from childhood cognitive ability, and finally, these disease events were included as time-varying covariates in dementia prediction models. To assess potential non-linear associations, we additionally fitted generalized additive models to a binary dementia outcome (ever vs never diagnosed during follow-up) using a logistic link and compared these models with corresponding linear logistic regression models.

*Statistical analyses of diseases other than dementia*

We defined time to first non-dementia condition as the interval from age at cognitive assessment to the earliest qualifying inpatient diagnosis. Participants without such diagnoses were censored at death or end of follow-up, consistent with the dementia models. Cox proportional hazards regression models with age as the underlying time scale were estimated using the same covariate specification as in the primary dementia analyses (standardized childhood cognitive test scores, parental education, and sex).

**Supplementary Results**

**Table S5. Association between childhood cognitive performance and dementia, main model (n = 10,539, 287 cases).**

| **Variable** | **Hazard Ratio** | **95% CI** | **P Value** |
| --- | --- | --- | --- |
| Inductive reasoning (per SD) | 0.84 | 0.72–0.98 | .022 |
| Spatial ability (per SD) | 0.99 | 0.86–1.13 | .830 |
| Verbal ability (per SD) | 1.00 | 0.86–1.15 | .981 |
| Parental education (per SD) | 1.04 | 0.93–1.17 | .491 |
| Sex (female vs male) | 0.79 | 0.62–0.99 | .043 |

**Table S6. Association between childhood cognitive performance and dementia, main model, adjusted for individual educational attainment in midlife (n = 9,640, 280 cases).**

| **Variable** | **Hazard Ratio** | **95% CI** | **P Value** |
| --- | --- | --- | --- |
| Inductive reasoning (per SD) | 0.83 | 0.71–0.96 | .015 |
| Spatial ability (per SD) | 1.02 | 0.89–1.17 | .754 |
| Verbal ability (per SD) | 1.00 | 0.86–1.16 | .982 |
| Parental education (per SD) | 1.07 | 0.95–1.21 | .263 |
| Midlife education (per SD) | 0.98 | 0.85–1.13 | .759 |
| Sex (female vs male) | 0.79 | 0.62–1.00 | .049 |

*Sensitivity Analyses*

*Post-hoc tests of isolated subtests to address potential partialling problem*

As a post-hoc follow-up test, to examine whether the domain-specific pattern of results reflected an artefact of jointly modelling correlated subtests, each cognitive domain was entered in a separate model alongside parental education and sex. When modelled alone, inductive reasoning was significantly associated with lower dementia risk (HR per SD = 0.83, 95% CI 0.74–0.94). Spatial ability (HR 0.91, 95% CI 0.81–1.02) and verbal ability (HR 0.90, 95% CI 0.80–1.02) were not significantly associated with dementia when entered alone. This pattern closely replicates the joint model, indicating that the specificity of the inductive reasoning association is not attributable to statistical partialling.

*Subtest reliability correction*

Because the inductive reasoning subtest had slightly higher reliability (0.92) than the verbal (0.87) and spatial (0.88) subtests, we conducted sensitivity analyses correcting for attenuation due to measurement error. Standardized test scores were divided by the square root of their respective reliability coefficients and re-standardized prior to analysis. Cox models using reliability-corrected scores, with parental education and sex as covariates, yielded results virtually identical to the primary analyses. Inductive reasoning remained significantly associated with dementia risk (HR, 0.84; 95% CI, 0.72–0.98), whereas spatial (HR, 0.99, 95% CI, 0.86–1.13) and verbal (HR, 1.00, 95% CI, 0.93–1.15) abilities were not associated. These findings indicate that small differences in reliability cannot account for the observed domain-specific pattern.

*Sensitivity to birth month*

To assess whether birth month within the sampling year influenced results, given that children born earlier in the year are older at the time of testing, birth month was added as a standardized covariate in the primary dementia model. The association between inductive reasoning and dementia was virtually unchanged (HR 0.84, 95% CI 0.73–0.98), and birth month itself was not associated with dementia risk (HR 0.93, 95% CI 0.83–1.05), indicating that relative age at testing did not confound the findings.

*Analysis of possible loss to follow-up by migration*

Recorded emigration does not necessarily equal loss to follow-up in Swedish registry data, e.g., as CDR aims to also include cases abroad.(6) In a sensitivity analysis excluding 262 participants whose last recorded status was emigration with no subsequent death recorded in the CDR, results were materially unchanged (inductive reasoning HR 0.84, 95% CI 0.73–0.98, n = 10,308, 287 dementia events).

*Analysis of competing risk*

Because lower childhood cognitive ability has been consistently associated with premature mortality in prior studies,(7, 8) differential mortality may introduce competing risk in dementia analyses. We therefore conducted competing risk analyses using the Fine–Gray sub-distribution hazards model treating death before dementia as a competing event.(9) Results were materially unchanged (sub-distribution HR, 0.87; 95% CI, 0.77–0.98), indicating that competing mortality did not explain the observed association.

**Table S7. Association between childhood cognitive performance and incident major somatic morbidity other than dementia (from diseases listed in the Charlson Comorbidity Index (CCI), n = 10,539, 5,183 cases).**

| **Variable** | **Hazard Ratio** | **95% CI** | **P Value** |
| --- | --- | --- | --- |
| Inductive reasoning (per SD) | 0.91 | 0.88–0.94 | <.001 |
| Spatial ability (per SD) | 0.99 | 0.96–1.02 | .604 |
| Verbal ability (per SD) | 0.98 | 0.95–1.02 | .270 |
| Parental education (per SD) | 0.94 | 0.91–0.97 | <.001 |
| Sex (female vs male) | 0.79 | 0.75–0.84 | <.001 |

**Table S8. Association between childhood cognitive performance and incident major somatic morbidity other than dementia (from diseases listed in the CCI) when adjusting also for midlife education (n = 9,640, 4,852 cases).**

| **Variable** | **Hazard Ratio** | **95% CI** | **P Value** |
| --- | --- | --- | --- |
| Inductive reasoning (per SD) | 0.92 | 0.89–0.96 | <.001 |
| Spatial ability (per SD) | 1.00 | 0.97–1.04 | .799 |
| Verbal ability (per SD) | 1.02 | 0.99–1.06 | .192 |
| Parental education (per SD) | 0.96 | 0.94–1.00 | .064 |
| Midlife education (per SD) | 0.91 | 0.88–0.94 | <.001 |
| Sex (female vs male) | 0.80 | 0.76–0.85 | <.001 |

**Table S9. Dementia Model Adjusted for Major Somatic Morbidity (CCI) (n = 10,539, 287 dementia cases).**

| **Variable** | **Hazard Ratio** | **95% CI** | **P Value** |
| --- | --- | --- | --- |
| Inductive reasoning (per SD) | 0.85 | 0.73–0.99 | .032 |
| Spatial ability (per SD) | 0.98 | 0.86–1.13 | .819 |
| Verbal ability (per SD) | 1.00 | 0.86–1.16 | .993 |
| Parental education (per SD) | 1.05 | 0.93–1.18 | .418 |
| Sex (female vs male) | 0.81 | 0.64–1.03 | .087 |
| Somatic morbidity | 1.63 | 1.28–2.07 | <.001 |

**Table S10. Dementia Model Adjusted for Major Somatic Morbidity (CCI) and midlife education (n = 9640, 280 cases).**

| **Variable** | **Hazard Ratio** | **95% CI** | **P Value** |
| --- | --- | --- | --- |
| Inductive reasoning (per SD) | 0.84 | 0.72–0.97 | .021 |
| Spatial ability (per SD) | 1.02 | 0.89–1.17 | .778 |
| Verbal ability (per SD) | 1.00 | 0.85–1.16 | .951 |
| Parental education (per SD) | 1.08 | 0.95–1.21 | .242 |
| Midlife education (per SD) | 0.99 | 0.86–1.14 | .871 |
| Sex (female vs male) | 0.81 | 0.64–1.03 | .090 |
| Somatic morbidity | 1.58 | 1.24–2.01 | <.001 |

**Table S11. Association Between Childhood Cognitive Performance and Incident Cerebrovascular Disease (n = 10,539, 1,208 cases).**

| **Variable** | **Hazard Ratio** | **95% CI** | **P Value** |
| --- | --- | --- | --- |
| Inductive reasoning (per SD) | 0.91 | 0.85–0.98 | .011 |
| Spatial ability (per SD) | 1.06 | 0.99–1.13 | .087 |
| Verbal ability (per SD) | 0.95 | 0.89–1.02 | .170 |
| Parental education (per SD) | 0.98 | 0.92–1.04 | .530 |
| Sex (female vs male) | 0.74 | 0.66–0.83 | <.001 |

**Table S12. Association Between Childhood Cognitive Performance and Dementia Risk Adjusted for Incident Cerebrovascular Disease (n = 10,539, 287 dementia cases).**

| **Variable** | **Hazard Ratio** | **95% CI** | **P Value** |
| --- | --- | --- | --- |
| Inductive reasoning (per SD) | 0.85 | 0.73–0.98 | .026 |
| Spatial ability (per SD) | 0.98 | 0.85–1.12 | .754 |
| Verbal ability (per SD) | 1.01 | 0.87–1.16 | .944 |
| Parental education (per SD) | 1.04 | 0.93–1.17 | .465 |
| Sex (female vs male) | 0.81 | 0.64–1.02 | .074 |
| Incident cerebrovascular disease | 2.38 | 1.73–3.28 | <.001 |

**Table S13. Association Between Childhood Cognitive Performance and Incident Myocardial Infarction (n = 10,539, 913 cases)**

| **Variable** | **Hazard Ratio** | **95% CI** | **P Value** |
| --- | --- | --- | --- |
| Inductive reasoning (per SD) | 0.82 | 0.76–0.89 | <.001 |
| Spatial ability (per SD) | 1.01 | 0.94–1.09 | .768 |
| Verbal ability (per SD) | 1.01 | 0.93–1.09 | .880 |
| Parental education (per SD) | 0.96 | 0.89–1.03 | .247 |
| Sex (female vs male) | 0.36 | 0.31–0.42 | <.001 |

**Table S14. Association Between Childhood Cognitive Performance and Dementia Risk Adjusted for Incident Myocardial Infarction (n = 10,539, 287 dementia cases).**

| **Variable** | **Hazard Ratio** | **95% CI** | **P Value** |
| --- | --- | --- | --- |
| Inductive reasoning (per SD) | 0.84 | 0.73–0.98 | .024 |
| Spatial ability (per SD) | 0.98 | 0.86–1.13 | .826 |
| Verbal ability (per SD) | 1.00 | 0.86–1.15 | .972 |
| Parental education (per SD) | 1.04 | 0.93–1.17 | .479 |
| Sex (female vs male) | 0.80 | 0.63–1.01 | .065 |
| Incident Myocardial infarction | 1.31 | 0.86–2.00 | .213 |

**Table S15. Association Between Childhood Cognitive Performance and Incident Heart Failure (n = 10,539, 819 cases).**

| **Variable** | **Hazard Ratio** | **95% CI** | **P Value** |
| --- | --- | --- | --- |
| Inductive reasoning (per SD) | 0.80 | 0.73–0.87 | <.001 |
| Spatial ability (per SD) | 1.05 | 0.97–1.13 | .256 |
| Verbal ability (per SD) | 1.00 | 0.91–1.09 | .942 |
| Parental education (per SD) | 0.87 | 0.80–0.95 | .002 |
| Sex (female vs male) | 0.59 | 0.51–0.68 | <.001 |

**Table S16. Association Between Childhood Cognitive Performance and Dementia Adjusted for Incident Heart Failure (n = 10,539, 287 dementia cases).**

| **Variable** | **Hazard Ratio** | **95% CI** | **P Value** |
| --- | --- | --- | --- |
| Inductive reasoning (per SD) | 0.85 | 0.73–0.98 | .029 |
| Spatial ability (per SD) | 0.98 | 0.86–1.12 | .793 |
| Verbal ability (per SD) | 1.00 | 0.86–1.15 | .980 |
| Parental education (per SD) | 1.05 | 0.93–1.18 | .442 |
| Sex (female vs male) | 0.80 | 0.64–1.02 | .070 |
| Incident heart failure | 2.15 | 1.40–3.31 | <.001 |

**Table S17. Association Between Childhood Cognitive Subtests and Incident Any Cardiovascular Disease (n = 10,539, 2,367 cases).**

| **Variable** | **Hazard Ratio** | **95% CI** | **P Value** |
| --- | --- | --- | --- |
| Inductive reasoning (per SD) | 0.88 | 0.83–0.92 | <.001 |
| Spatial ability (per SD) | 1.02 | 0.98–1.07 | .347 |
| Verbal ability (per SD) | 0.97 | 0.92–1.02 | .274 |
| Parental education (per SD) | 0.95 | 0.91–0.99 | .029 |
| Sex (female vs male) | 0.56 | 0.51–0.61 | <.001 |

**Table S18. Association Between Childhood Cognitive Subtests and Incident Any Cardiovascular Disease adjusted also for midlife education (n = 9,640, 2,220 cases).**

| **Variable** | **Hazard Ratio** | **95% CI** | **P Value** |
| --- | --- | --- | --- |
| Inductive reasoning (per SD) | 0.89 | 0.85–0.94 | <.001 |
| Spatial ability (per SD) | 1.03 | 0.98–1.08 | .220 |
| Verbal ability (per SD) | 1.01 | 0.96–1.07 | .618 |
| Parental education (per SD) | 0.98 | 0.94–1.03 | .488 |
| Midlife education (per SD) | 0.91 | 0.86–0.96 | <.001 |
| Sex (female vs male) | 0.56 | 0.51–0.61 | <.001 |

**Table S19. Dementia risk adjusted for incident any cardiovascular disease (n = 10,539, 287 dementia cases).**

| **Variable** | **Hazard Ratio** | **95% CI** | **P Value** |
| --- | --- | --- | --- |
| Inductive reasoning (per SD) | 0.85 | 0.73–0.99 | .032 |
| Spatial ability (per SD) | 0.98 | 0.85–1.12 | .770 |
| Verbal ability (per SD) | 1.00 | 0.87–1.16 | .981 |
| Parental education (per SD) | 1.05 | 0.93–1.18 | .419 |
| Sex (female vs male) | 0.85 | 0.67–1.07 | .170 |
| Incident any CVD | 2.00 | 1.53–2.63 | <.001 |

**Table S20. Dementia risk adjusted for incident any cardiovascular disease and midlife education (n = 9,640, 280 dementia cases)**

| **Variable** | **Hazard Ratio** | **95% CI** | **P Value** |
| --- | --- | --- | --- |
| Inductive reasoning (per SD) | 0.84 | 0.72–0.97 | .021 |
| Spatial ability (per SD) | 1.02 | 0.89–1.17 | .817 |
| Verbal ability (per SD) | 1.00 | 0.85–1.16 | .969 |
| Parental education (per SD) | 1.08 | 0.95–1.21 | .241 |
| Midlife education (per SD) | 0.99 | 0.86–1.14 | .865 |
| Sex (female vs male) | 0.85 | 0.67–1.08 | .181 |
| Incident any CVD (time-varying) | 1.99 | 1.74–2.84 | <.001 |

**Table S21. Association Between Childhood Cognitive Subtests and Incident Diabetes (n = 10,539, 308 cases).**

| **Variable** | **Hazard Ratio** | **95% CI** | **P Value** |
| --- | --- | --- | --- |
| Inductive reasoning (per SD) | 0.74 | 0.64–0.86 | <.001 |
| Spatial ability (per SD) | 0.98 | 0.86–1.12 | .757 |
| Verbal ability (per SD) | 0.98 | 0.85–1.13 | .772 |
| Parental education (per SD) | 0.84 | 0.72–0.98 | .030 |
| Sex (female vs male) | 0.54 | 0.43–0.68 | <.001 |

**Table S22. Association Between Childhood Cognitive Subtests and Incident Diabetes, adjusted also for midlife education (n = 9640, 282 cases).**

| **Variable** | **Hazard Ratio** | **95% CI** | **P Value** |
| --- | --- | --- | --- |
| Inductive reasoning (per SD) | 0.75 | 0.64–0.87 | <.001 |
| Spatial ability (per SD) | 0.99 | 0.87–1.14 | .904 |
| Verbal ability (per SD) | 1.05 | 0.90–1.22 | .543 |
| Parental education (per SD) | 0.90 | 0.76–1.06 | .200 |
| Midlife education (per SD) | 0.84 | 0.73–0.97 | .016 |
| Sex (female vs male) | 0.51 | 0.40–0.66 | <.001 |

**Table S23. Dementia Risk Adjusted for Diabetes (n = 10,539, 287 cases).**

| **Variable** | **Hazard Ratio** | **95% CI** | **P Value** |
| --- | --- | --- | --- |
| Inductive reasoning (per SD) | 0.85 | 0.73–0.98 | .028 |
| Spatial ability (per SD) | 0.99 | 0.86–1.13 | .832 |
| Verbal ability (per SD) | 1.00 | 0.86–1.15 | .962 |
| Parental education (per SD) | 1.04 | 0.93–1.17 | .472 |
| Sex (female vs male) | 0.79 | 0.63–1.00 | .053 |
| Incident diabetes (time-varying) | 2.74 | 1.53–4.88 | <.001 |

**Table S24. Dementia Risk Adjusted for Diabetes and for midlife education (n = 9640, 280 cases).**

| **Variable** | **Hazard Ratio** | **95% CI** | **P Value** |
| --- | --- | --- | --- |
| Inductive reasoning (per SD) | 0.83 | 0.72–0.97 | .019 |
| Spatial ability (per SD) | 1.02 | 0.89–1.17 | .743 |
| Verbal ability (per SD) | 1.00 | 0.85–1.16 | .952 |
| Parental education (per SD) | 1.07 | 0.95–1.21 | .258 |
| Midlife education (per SD) | 0.98 | 0.85–1.13 | .791 |
| Sex (female vs male) | 0.80 | 0.63–1.01 | .062 |
| Incident diabetes (time-varying) | 2.76 | 1.54–4.92 | <.001 |

*Details on analysis results in the expanded cohort*

To evaluate robustness of results, we added the UGU birth 1953 cohort who completed the same cognitive tests in 6^th^ grade. Given their younger age at follow-up and fewer dementia cases, we were not powered to analyze this cohort separately. In addition, the parental education measures were not recorded the same way for this cohort. Thus, all individuals with complete data on inductive reasoning and sex were included here: 10,547 persons in the 1948 cohort, of whom 288 with a dementia diagnosis (the additional 8 persons lacked any measure of parental education preventing inclusion in the main analytic sample, the additional dementia case here was identified both in NPR and CDR), and 9372 persons in the 1953 cohort, 79 (0.84%) with a dementia diagnosis (of which 8 identified only in CDR, 46 only in NPR, and 25 in both). This yielded a total of 19,919 individuals (10,100 males) and 367 dementia cases (1.84%). When estimating the model using the combined cohorts with inductive reasoning score and sex as predictors, inductive reasoning was associated with lower hazard of dementia with comparable effect size (HR = 0.85, 95% CI: 0.77–0.95, p = 0.003) and sex was also associated, with women having lower dementia risk (HR = 0.802, 95% CI: 0.65–0.99).

To assess potential non-linear associations (10), we fitted a generalized additive model with a penalized spline in the expanded sample, using raw reasoning scores. The smooth term was significant (edf = 1.389, χ² = 6.683, p = .016), but the effective degrees of freedom were close to 1, indicating that the relationship was essentially linear (Figure 3). A formal comparison between the GAM (using the R package mgcv) and a simple logistic regression model revealed no improvement in fit for the non-linear specification (ΔDeviance = 1.006, p = .117). These results show that the association between childhood inductive reasoning and dementia risk is well-described by a linear effect.
